# Reduced Odds of SARS-CoV-2 Reinfection after Vaccination among New York City Adults, June–August 2021

**DOI:** 10.1101/2021.12.09.21267203

**Authors:** Alison Levin-Rector, Lauren Firestein, Emily McGibbon, Jessica Sell, Sungwoo Lim, Ellen H. Lee, Don Weiss, Anita Geevarughese, Jane R. Zucker, Sharon K. Greene

## Abstract

**Background:** Belief in immunity from prior infection and concern that vaccines might not protect against new variants are contributors to vaccine hesitancy. We assessed effectiveness of full and partial COVID-19 vaccination against reinfection when Delta was the predominant variant in New York City.

**Methods:** We conducted a case-control study in which case-patients with reinfection during June 15– August 31, 2021 and control subjects with no reinfection were matched (1:3) on age, sex, timing of initial positive test in 2020, and neighborhood poverty level. Conditional logistic regression was used to calculate matched odds ratios (mOR) and 95% confidence intervals (CI).

**Results:** Of 349,598 adult residents who tested positive for SARS-CoV-2 infection in 2020, did not test positive again >90 days after initial positive test through June 15, 2021, and did not die before June 15, 2021, 1,067 were reinfected during June 15–August 31, 2021. Of 1,048 with complete matching criteria data, 499 (47.6%) were known to be symptomatic for COVID-19-like-illness, and 75 (7.2%) were hospitalized. Unvaccinated individuals, compared with fully vaccinated individuals, had elevated odds of reinfection (mOR, 2.23; 95% CI, 1.90, 2.61), of symptomatic reinfection (mOR, 2.17; 95% CI, 1.72, 2.74), and of reinfection with hospitalization (mOR, 2.59; 95% CI, 1.43, 4.69). Partially versus fully vaccinated individuals had 1.58 (95% CI: 1.22, 2.06) times the odds of reinfection. All three vaccines authorized or approved for use in the U.S. were similarly effective.

**Conclusion:** Among adults with previous SARS-CoV-2 infection, vaccination reduced odds of reinfections when the Delta variant predominated.

## Introduction

The first cases of severe acute respiratory syndrome coronavirus 2 (SARS-CoV-2) causing coronavirus disease 2019 (COVID-19) were detected in New York City (NYC) in February 2020 [1, 2]. NYC quickly became a pandemic epicenter, with daily case counts exceeding 6,000 [2]. By December 31, 2020, 433,478 individuals had tested positive for SARS-CoV-2 infection among approximately 7.1 million adult NYC residents [2], and an unknown number of infected persons were never tested due to inability to access care and limited initial testing availability [2, 3].

On December 14, 2020, a NYC health care worker received the first COVID-19 vaccine dose administered under Phase 1 of New York State’s Vaccine Distribution Plan [4]. Subsequently, eligibility and availability in NYC expanded for the three COVID-19 vaccines authorized or approved in the U.S., i.e., BNT162b2 from Pfizer-BioNTech, mRNA-1273 from Moderna, and Ad26.COV2 from Janssen (Johnson & Johnson); all three vaccines are highly effective at preventing SARS-CoV-2 infection and severe outcomes, including hospitalization and death, and serious adverse events after vaccination are rare [5-11].

Increasing vaccination rates became the core strategy to enable NYC’s reopening, including mandates for health care workers [12] and City employees [13], and the “Key to NYC” program requiring vaccination proof for indoor activities, such as dining and entertainment [14]. Despite strong evidence of COVID-19 vaccine safety and effectiveness and vaccination mandates, 15.1% of NYC residents ≥18 years-old were not vaccinated as of October 19, 2021 [2]. A national survey conducted in June 2020 before any COVID-19 vaccine was available found that 12% of respondents who anticipated rejecting vaccination cited the belief that they were already immune from prior infection [15]. According to the NYC Health Opinion Poll in June 2021, of NYC adults who were unvaccinated and did not intend to or were unsure if they would get vaccinated in the future, one-quarter reported that they did not think they needed a COVID-19 vaccine because of immunity from prior infection (Sarah Dumas, NYC Department of Health and Mental Hygiene [DOHMH], unpublished data,^1^ 2021). Half were concerned that COVID-19 vaccines might not protect people against new variants (ibid.).

The B.1.617.2 variant, also known as Delta, was classified as a variant of concern by the World Health Organization (WHO) in May 2021 [16]. Given its high transmissibility, Delta constituted approximately a third of sequenced cases among NYC residents as of mid-June 2021, half of sequenced cases as of early July, and nearly all (97%) by late July [2]. Modest reductions in COVID-19 vaccine effectiveness against symptomatic disease with the Delta variant as compared with the Alpha variant have been noted [9, 17, 18].

Cavanaugh et al. conducted a case-control study of the association between vaccination and SARS-CoV-2 reinfection in Kentucky during May–June 2021 among persons who previously tested positive for SARS-CoV-2 infection in 2020. Kentucky residents who were unvaccinated had 2.34 times the odds of reinfection compared with those who were fully vaccinated, supporting recommendations for COVID-19 vaccination for those with prior SARS-CoV-2 infection [19]. In Health and Human Services Region 4, which includes Kentucky, Delta constituted a minority of sequenced cases through June 2021 [20]. As this study was conducted in a predominantly rural state and before Delta predominance, it might have limited generalizability. Vaccine effectiveness against reinfection in an urban population while Delta is the dominant variant is unknown.

We aimed to assess vaccine effectiveness against reinfection among NYC adult residents who tested positive for SARS-CoV-2 infection in 2020. We assessed effectiveness of full and partial COVID-19 vaccination, overall and by vaccine manufacturer, against the outcomes of reinfection, symptomatic reinfection, and reinfection with hospitalization during June 15–August 31, 2021, when the Delta variant predominated.

## Methods

### Study Population and Data Source

The study population consisted of NYC residents who tested positive for SARS-CoV-2 by a molecular or antigen test in 2020 and did not have evidence of reinfection, defined as a positive SARS-CoV-2 molecular or antigen test result >90 days after the initial positive test, before June 15, 2021. We excluded residents of selected congregate settings (i.e., nursing homes, adult care facilities, and jails), individuals <18-years-old as of their initial positive test in 2020, and individuals who died before June 15, 2021.

Demographic, laboratory, hospitalization, and mortality data were collected as part of routine public health surveillance and vital statistics monitoring by DOHMH, as previously described [1]. Symptom status was ascertained by routine interview, e.g., for contact tracing. Data were extracted from the DOHMH COVID-19 surveillance database on September 10, 2021.

### Cases and Controls

To assess vaccine effectiveness against reinfection, we defined case-patients as individuals who tested positive for SARS-CoV-2 reinfection during June 15–August 31, 2021. In a subset analysis, we restricted to case-patients with symptomatic reinfection to reduce bias from differential ascertainment of asymptomatic infections among populations with frequent testing (e.g., as an occupational requirement), which could vary by vaccination status. Symptomatic reinfections were defined as case-patients who met the clinical criteria for COVID-19-like illness, per the U.S. Centers for Disease Control and Prevention and Council of State and Territorial Epidemiologists case definition [21]. Control subjects were selected from the study population as having had no documented reinfection through August 31, 2021.

To assess protection by vaccination against reinfection resulting in severe illness, we further defined case-patients as individuals who tested positive for SARS-CoV-2 reinfection during June 15–August 31, 2021 and who met criteria for COVID-19 hospitalization. COVID-19 hospitalization was defined as a hospitalization within +/- 14 days of a positive SARS-CoV-2 test or at time of COVID-19 death (which was defined as death within +/- 30 days of a positive SARS-CoV-2 test or where COVID-19 was listed as a cause of death on the death certificate).

Control subjects for this analysis were selected from the study population as having had either no reinfection through August 31, 2021 or a reinfection during June 15–August 31, 2021 but no hospitalization or death. We did not separately assess protection by vaccination against reinfection resulting in death because only two individuals from the study population who tested positive for SARS-CoV-2 reinfection during June 15–August 31, 2021 died.

Three control subjects were matched to each case-patient on sex, age within +/- 3 years, specimen collection date in 2020 of initial positive SARS-CoV-2 test within +/- 1 week, and neighborhood poverty level. Neighborhood poverty (based on census tract of residence as of initial laboratory report in 2020) was defined as the percent of residents with incomes below the federal poverty level, per American Community Survey 2014–2018 [22]. The controls that matched most closely on age and specimen collection date were selected.

### Exposure

By matching with the DOHMH Citywide Immunization Registry [23], case-patients were assigned a vaccination status based on their reinfection date, defined as the specimen collection date of the first positive test indicating reinfection, and control subjects were assigned a vaccination status based on the reinfection date of their matched case-patient. Individuals were considered fully vaccinated if they received two doses of a COVID-19 mRNA vaccine (Pfizer-BioNTech or Moderna) or one dose of a viral vector vaccine (Janssen) ≥14 days before the reinfection date. Partially vaccinated individuals received one dose of an mRNA vaccine ≥14 days before the reinfection date, and unvaccinated individuals did not receive any vaccine doses ≥14 days before the reinfection date.

### Analysis

Matched odds ratios (mORs) and 95% confidence intervals (CIs) were calculated using conditional logistic regression to estimate the odds of reinfection, of symptomatic reinfection, and of reinfection with hospitalization by vaccination status, with fully vaccinated persons as the referent. Odds of reinfection by vaccine manufacturer were also estimated. In addition, vaccine effectiveness (VE) was calculated as 100% × (1 – mOR), with unvaccinated persons as the referent [24]. Case-patients with identical matching criteria and their matched controls were pooled to retain more data and increase precision. A sensitivity analysis was performed to assess potential selection and confounding bias by using a case-control weighted target maximum likelihood estimation method (CCW-TMLE) (Supplementary Material). Statistical analyses were performed using SAS Enterprise Guide, version 7.1 (SAS Institute). This work was deemed public health surveillance that is non-research by the DOHMH Institutional Review Board.

## Results

Of 432,360 NYC residents who tested positive for SARS-CoV-2 infection in 2020, 349,598 (80.9%) were eligible for analysis of COVID-19 vaccination status and reinfection (Figure). Of these, 1,067 (0.3%) were reinfected during June 15–August 31, 2021. These 1,067 individuals represented 1.5% of the 69,094 NYC adults who did not reside in congregate settings and tested positive for SARS-CoV-2 infection during this period. Of the 1,067, 1,048 (98.2%) had complete data on matching criteria and were included in analysis, and of these, 499 (47.6%) were symptomatic for COVID-19-like illness. Of the remaining 549, 228 (21.8%) identified as asymptomatic, 265 (25.3%) had missing symptom information, and 56 (5.3%) had symptoms that did not meet clinical criteria for COVID-19-like-illness. Seventy-five (7.2%) case-patients were hospitalized. Case-patients and matched control subjects were similar on sex, age, timing of initial positive test in 2020, and neighborhood poverty level (Table 1). Reinfections were most common among females, case-patients aged 25–34 years, and case-patients who first tested positive for SARS-CoV-2 infection in April or December 2020. Reinfections and symptomatic reinfections were most common among residents of neighborhoods of low and medium poverty levels, while hospitalized case-patients with reinfection most commonly resided in medium and high poverty neighborhoods (Table 1).

**Figure.**
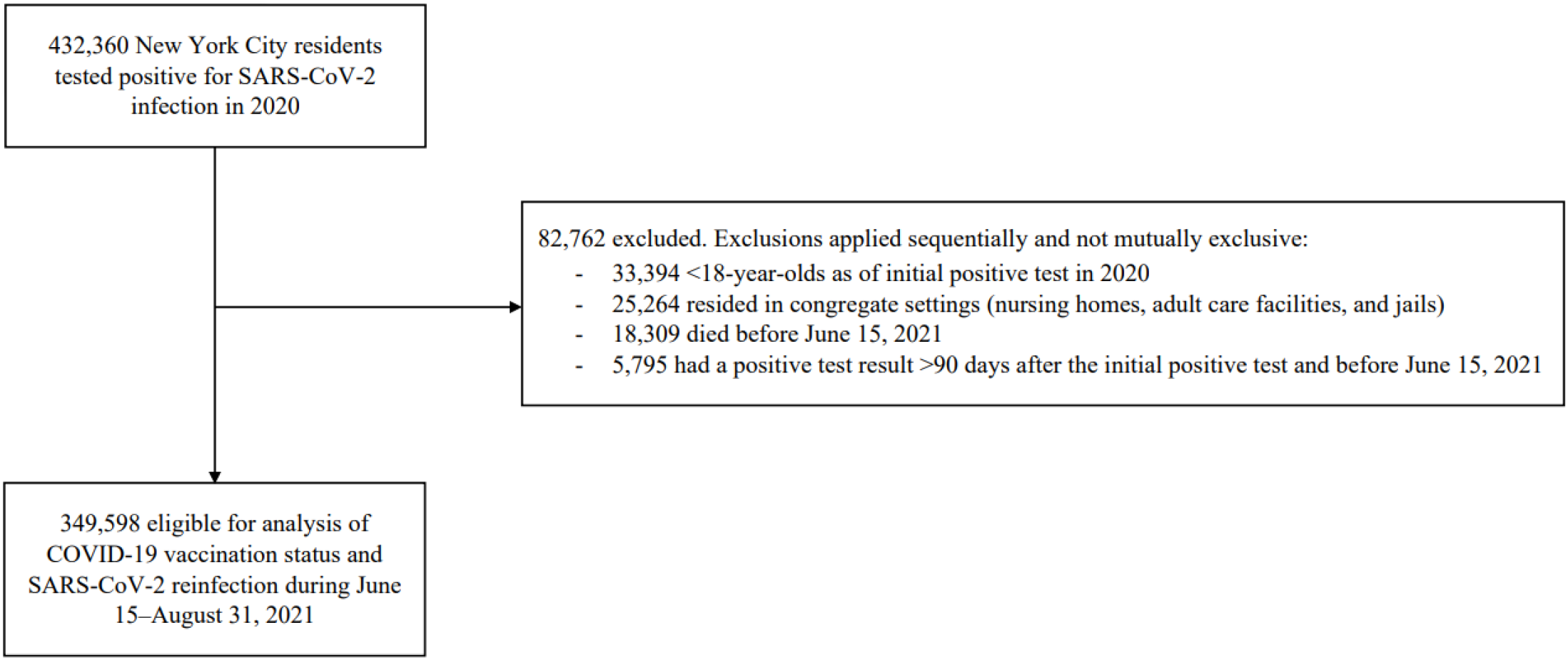
Eligibility for analysis of COVID-19 vaccination status and SARS-CoV-2 reinfection during June 15–August 31, 2021 among New York City residents with first SARS-CoV-2 infection in 2020

**Table 1.** Characteristics of case-patients with SARS-CoV-2 reinfection during June 15–August 31, 2021 and matched control subjects without reinfection among New York City adults with first SARS-CoV-2 infection in 2020

|  | **Reinfection vs. no reinfection** |  | **Symptomatic reinfection vs. no reinfection** |  | **Reinfection with hospitalization vs. no reinfection<sup>1</sup>** |  |
| --- | --- | --- | --- | --- | --- | --- |
|  | Cases<br>N (%) | Controls<br>N (%) | Cases<br>N (%) | Controls<br>N (%) | Cases<br>N (%) | Controls<br>N (%) |
| **Total** | 1,048 | 3,144 | 499 | 1,497 | 75 | 225 |
| **Sex** |  |  |  |  |  |  |
| **Female** | 603 (57.5%) | 1,809 (57.5%) | 295 (59.1%) | 885 (59.1%) | 39 (52.0%) | 117 (52.0%) |
| **Age group<sup>2</sup>** |  |  |  |  |  |  |
| **18-24** | 148 (14.1%) | 449 (14.3%) | 73 (14.6%) | 220 (14.7%) | 4 (5.3%) | 12 (5.3%) |
| **25-34** | 375 (35.8%) | 1,121 (35.7%) | 190 (38.1%) | 569 (38.0%) | 17 (22.7%) | 51 (22.7%) |
| **35-44** | 203 (19.4%) | 604 (19.2%) | 102 (20.4%) | 306 (20.4%) | 11 (14.7%) | 33 (14.7%) |
| **45-54** | 130 (12.4%) | 391 (12.4%) | 57 (11.4%) | 168 (11.2%) | 11 (14.7%) | 34 (15.1%) |
| **55-64** | 109 (10.4%) | 331 (10.5%) | 47 (9.4%) | 146 (9.8%) | 13 (17.3%) | 38 (16.9%) |
| **65-74** | 53 (5.1%) | 160 (5.1%) | 19 (3.8%) | 55 (3.7%) | 13 (17.3%) | 39 (17.3%) |
| **75+** | 30 (2.9%) | 88 (2.8%) | 11 (2.2%) | 33 (2.2%) | 6 (8.0%) | 18 (8.0%) |
| **Census tract-based poverty level<sup>2</sup>** |  |  |  |  |  |  |
| **Low** | 339 (32.3%) | 1,017 (32.3%) | 168 (33.7%) | 504 (33.7%) | 13 (17.3%) | 39 (17.3%) |
| **Medium** | 328 (31.3%) | 984 (31.3%) | 153 (30.7%) | 459 (30.7%) | 28 (37.3%) | 84 (37.3%) |
| **High** | 220 (21.0%) | 660 (21.0%) | 101 (20.2%) | 303 (20.2%) | 21 (28.0%) | 63 (28.0%) |
| **Very high** | 161 (15.4%) | 483 (15.4%) | 77 (15.4%) | 231 (15.4%) | 13 (17.3%) | 39 (17.3%) |
| **Month of initial positive SARS-CoV-2 test (2020)** |  |  |  |  |  |  |
| **March** | 151 (14.4%) | 455 (14.5%) | 72 (14.4%) | 217 (14.5%) | 9 (12.0%) | 27 (12.0%) |
| **April** | 233 (22.2%) | 698 (22.2%) | 111 (22.2%) | 333 (22.2%) | 20 (26.7%) | 60 (26.7%) |
| **May** | 50 (4.8%) | 149 (4.7%) | 22 (4.4%) | 65 (4.3%) | 3 (4.0%) | 9 (4.0%) |
| **June** | 17 (1.6%) | 52 (1.7%) | 8 (1.6%) | 24 (1.6%) | 0 (0.0%) | 0 (0.0%) |
| **July** | 37 (3.5%) | 110 (3.5%) | 18 (3.6%) | 54 (3.6%) | 2 (2.7%) | 6 (2.7%) |
| **August** | 19 (1.8%) | 56 (1.8%) | 15 (3.0%) | 45 (3.0%) | 3 (4.0%) | 9 (4.0%) |
| **September** | 37 (3.5%) | 112 (3.6%) | 13 (2.6%) | 40 (2.7%) | 1 (1.3%) | 3 (1.3%) |
| **October** | 42 (4.0%) | 126 (4.0%) | 20 (4.0%) | 58 (3.9%) | 2 (2.7%) | 6 (2.7%) |
| **November** | 150 (14.3%) | 450 (14.3%) | 70 (14.0%) | 211 (14.1%) | 8 (10.7%) | 24 (10.7%) |
| **December** | 312 (29.8%) | 936 (29.8%) | 150 (30.1%) | 450 (30.1%) | 27 (36.0%) | 81 (36.0%) |
| **Vaccination Status<sup>3</sup>** |  |  |  |  |  |  |
| **Not vaccinated<sup>4</sup>** | 638 (60.9%) | 1,391 (44.2%) | 302 (60.5%) | 655 (43.8%) | 41 (54.7%) | 75 (33.3%) |
| **Partially vaccinated** | 96 (9.2%) | 291 (9.3%) | 43 (8.6%) | 149 (10.0%) | 7 (9.3%) | 29 (12.9%) |
| **Fully vaccinated** | 314 (30.0%) | 1,462 (46.5%) | 154 (30.9%) | 693 (46.3%) | 27 (36%) | 121 (53.8%) |
| **Vaccine manufacturer<sup>5</sup>** |  |  |  |  |  |  |
| **Not vaccinated** | 638 (67.0%) | 1,391 (48.8%) | 302 (66.2%) | 655 (48.6%) | 41 (60.3%) | 75 (38.3%) |
| **Fully vaccinated with Pfizer** | 174 (18.3%) | 824 (28.9%) | 91 (20.0%) | 390 (28.9%) | 16 (23.5%) | 66 (33.7%) |
| **Fully vaccinated with Moderna** | 116 (12.2%) | 523 (18.3%) | 53 (11.6%) | 246 (18.2%) | 9 (13.2%) | 48 (24.5%) |
| **Fully vaccinated with Janssen** | 24 (2.5%) | 115 (4.0%) | 10 (2.2%) | 57 (4.2%) | 2 (2.9%) | 7 (3.6%) |
| **Days from fully vaccinated<sup>5</sup> to reference date<sup>6</sup>** |  |  |  |  |  |  |
| **0–60** | 50 (15.9%) | 330 (22.6%) | 17 (11.0%) | 160 (23.1%) | 4 (14.8%) | 32 (26.5%) |
| **61–120** | 149 (47.5%) | 710 (48.6%) | 78 (50.7%) | 338 (48.8%) | 17 (63.0%) | 65 (53.7%) |
| **121–180** | 89 (28.3%) | 351 (24.0%) | 40 (26.0%) | 162 (23.4%) | 6 (22.2%) | 21 (17.4%) |
| **>180** | 26 (8.3%) | 71 (4.9%) | 19 (12.3%) | 33 (4.8%) | 0 (0.0%) | 3 (2.5%) |
| **Month tested positive for SARS-CoV-2 reinfection (2021)** |  |  |  |  |  |  |
| **June 15–30** | 70 (6.7%) | - | 24 (4.8%) | - | 20 (26.7%) | - |
| **July** | 304 (29.0%) | - | 155 (31.1%) | - | 20 (26.7%) | - |
| **August** | 674 (64.3%) | - | 320 (64.1%) | - | 35 (46.7%) | - |

| **Severity of initial infection** |  |  |  |  |  |  |
| --- | --- | --- | --- | --- | --- | --- |
| **Hospitalized** | 78 (7.4%) | 276 (8.8%) | 36 (7.2%) | 120 (8.0%) | 23 (30.7%) | 37 (16.4%) |

<sup>1</sup>For this case definition, matched control subjects were selected from the study population as having had either no reinfection through August 31, 2021 or a reinfection during June 15–August 31, 2021 but no hospitalization or death. Ultimately, all selected control subjects had no reinfection through August 31, 2021.
<sup>2</sup>Age and census tract of residence as of initial positive SARS-CoV-2 test in 2020. Low poverty level defined as <10% of residents below the federal poverty level, medium as 10% to <20%, high as 20 to <30%, and very high as ≥30%.
<sup>3</sup>Individuals were considered fully vaccinated if they received two doses of a COVID-19 mRNA vaccine (Pfizer-BioNTech or Moderna) or one dose of a viral vector vaccine (Janssen) ≥14 days before the reinfection date. Partially vaccinated individuals received one dose of an mRNA vaccine ≥14 days before the reinfection date, and unvaccinated individuals did not receive any vaccine doses ≥14 days before the reinfection date.
<sup>4</sup>Some individuals classified as unvaccinated received one dose of vaccine <14 days before the reinfection date: 14 case-patients with reinfection and 26 control subjects; 7 case-patients with symptomatic reinfection and 15 control subjects; and 3 case-patient with reinfection with hospitalization and 2 control subjects. Some individuals classified as partially vaccinated received a second dose of an mRNA vaccine <14 days before the reinfection date: 14 case-patients with reinfection and 78 control subjects; 4 case-
patients with symptomatic reinfection and 39 control subjects; 2 case-patients with reinfection with hospitalization and 3 control subjects.
<sup>5</sup>Partially vaccinated persons excluded.
<sup>6</sup>For case-patients, the reference date was the specimen collection date of the first positive test indicating SARS-CoV-2 reinfection in 2021. For control subjects, the reference date was the reinfection date of their matched case-patient.

NYC adult residents who initially tested positive for SARS-CoV-2 infection in 2020 and remained unvaccinated had elevated odds of reinfection (mOR, 2.23; 95% CI, 1.90, 2.61), of symptomatic reinfection (mOR, 2.17; 95% CI, 1.72, 2.74), and of reinfection with hospitalization (mOR, 2.59; 95% CI, 1.43, 4.69) during June 15–August 31, 2021 compared with those who were fully vaccinated (Table 2). In other words, with unvaccinated persons as the referent, VE for full vaccination against reinfection was 55% (95% CI, 47%, 62%), against symptomatic reinfection was 54% (95% CI, 42%, 63%), and against reinfection with hospitalization was 61% (95% CI, 30%, 79%). Findings were robust to reanalysis using CCW-TMLE, suggesting negligible selection and confounding bias (Supplementary Material).

**Table 2.** Associations of SARS-CoV-2 reinfection during June 15–August 31, 2021 with COVID-19 vaccination status and vaccine manufacturer among New York City adults with first SARS-CoV-2 infection in 2020

|  | Reinfection vs.<br>no reinfection |  | Symptomatic reinfection<br>vs.<br>no reinfection |  | Reinfection with<br>hospitalization vs. no<br>reinfection<sup>1</sup> |  |
| --- | --- | --- | --- | --- | --- | --- |
|  | mOR (95% CI) | p-value | mOR (95% CI) | p-value | mOR (95% CI) | p-value |
| **Vaccination status** |  |  |  |  |  |  |
| **Fully vaccinated as referent** |  |  |  |  |  |  |
| Not vaccinated | 2.23 (1.90, 2.61) | <0.0001 | 2.17 (1.72, 2.74) | <0.0001 | 2.59 (1.43, 4.69) | 0.002 |
| Partially vaccinated | 1.58 (1.22, 2.06) | 0.001 | 1.34 (0.91, 1.91) | 0.14 | 1.13 (0.45, 2.82) | 0.80 |
| **Not vaccinated as referent** |  |  |  |  |  |  |
| Partially vaccinated | 0.71 (0.55, 0.91) | 0.008 | 0.62 (0.43, 0.89) | 0.01 | 0.44 (0.18, 1.08) | 0.07 |
| Fully vaccinated | 0.45 (0.38, 0.53) | <0.0001 | 0.46 (0.37, 0.58) | <0.0001 | 0.39 (0.21, 0.70) | 0.002 |
| **Vaccine manufacturer, not vaccinated as referent** |  |  |  |  |  |  |
| Fully vaccinated with Pfizer | 0.44 (0.36, 0.54) | <0.0001 | 0.49 (0.37, 0.65) | <0.0001 | 0.42 (0.20, 0.88) | 0.02 |
| Fully vaccinated with Moderna | 0.46 (0.37, 0.58) | <0.0001 | 0.45 (0.32, 0.64) | <0.0001 | 0.32 (0.14, 0.74) | 0.007 |
| Vaccinated with Janssen | 0.45 (0.29, 0.71) | 0.0006 | 0.36 (0.18, 0.73) | 0.004 | 0.49 (0.10, 2.44) | 0.39 |

mOR: matched odds ratio; CI: confidence interval
<sup>1</sup>For this case definition, matched control subjects were selected from the study population as having had either no reinfection through August 31, 2021 or a reinfection during June 15–August 31, 2021 but no hospitalization or death. Ultimately, all selected control subjects had no reinfection through August 31, 2021.

Few partially vaccinated persons were included in analysis (Table 1). Compared with NYC residents who tested positive for SARS-CoV-2 infection in 2020 and were fully vaccinated, those who were partially vaccinated had 1.58 (95% CI: 1.22, 2.06) times the odds of reinfection (Table 2), i.e., VE for partial vaccination against reinfection was 29% (95% CI, 9%, 45%). No associations were observed for symptomatic reinfection and for reinfection with hospitalization comparing partially versus fully vaccinated persons (Table 2). With unvaccinated persons as the referent, VE for partial vaccination against symptomatic reinfection was 38% (95% CI, 11%, 57%) and against reinfection with hospitalization was 57% (95% CI, −8%, 83%).

Vaccines from all three manufacturers reduced the odds of reinfection (mOR point estimates similar by manufacturer, range 0.44–0.46, i.e., VE ≈ 55%), of symptomatic reinfection (mOR point estimates range: 0.36–0.49, i.e., VE range 51%–64%), and of reinfection with hospitalization (mOR point estimates range: 0.32–0.49, i.e., VE range 51%–68%). Only two patients with reinfection with hospitalization had received the Janssen vaccine (Table 1), so a significant protective effect against this outcome for this vaccine could not be demonstrated (p=0.39), but significant protective effects at p≤0.02 were demonstrated for the other 8 combinations of outcomes and vaccine manufacturers (Table 2).

## Discussion

Among adult NYC residents who previously tested positive for SARS-CoV-2 infection in 2020, unvaccinated individuals, compared with those who were fully vaccinated, had 2.23 (95% CI: 1.90, 2.61) times the odds and partially vaccinated individuals had 1.58 (95% CI: 1.22, 2.06) times the odds of reinfection during a period when the Delta variant predominated (June 15– August 31, 2021). Our findings are consistent with Cavanaugh et al., who found that among Kentucky residents who tested positive for SARS-CoV-2 infection in 2020, unvaccinated individuals had 2.34 (95% CI: 1.58, 3.47) times the odds and partially vaccinated individuals had 1.56 (95% CI: 0.81, 3.01) times the odds of reinfection during a period before Delta variant predominance (May–June 2021) compared with those who were fully vaccinated [19]. We additionally found that unvaccinated individuals had significantly greater odds of both symptomatic reinfection and of reinfection with hospitalization. The consistency of these mORs between Kentucky before Delta variant predominance and NYC during Delta variant predominance, as well as for symptomatic reinfection and reinfection with hospitalization in NYC, supports existing recommendations to vaccinate persons with a prior COVID-19 diagnosis [25].

As previously noted, there were few observations among individuals who were previously infected and were partially vaccinated, limiting our conclusions for this population. The odds of symptomatic reinfection or reinfection with hospitalization were higher with partial vaccination compared with full vaccination but not statistically significantly higher. Importantly, the odds of symptomatic reinfection with partial vaccination were lower compared with no vaccination, supporting the importance of vaccination for individuals with prior infection; similar results were seen for reinfection with hospitalization, though this was not statistically significant.

Individuals in this population fully vaccinated with any of the three vaccines authorized or approved for full or emergency use in the U.S. were similarly protected against SARS-CoV-2 reinfection. These vaccines are known to be protective against SARS-CoV-2 infection, including against the Delta variant [5, 9], but prior literature on reinfections is sparse. Our finding of 61% (95% CI, 30%, 79%) effectiveness of full vaccination against reinfection with hospitalization among previously infected individuals was lower than prior estimates of >90% VE against infection with hospitalization among infection-naïve individuals. This difference in estimates could be attributable to different study populations, in that the benefit of vaccination would be expected to be greater for individuals without prior immunity than for previously infected individuals.

There are at least four limitations to this study. First, persons who were infected with SARS-CoV-2 in 2020 but who were not tested (e.g., due to limited testing availability) could not be included. Thus, our analysis of 1,048 first reinfections during June 15–August 31, 2021 among those initially infected in 2020 is an undercount, reducing sample size and precision of estimates. Because symptom status was missing for a quarter of reinfected individuals, the number of symptomatic reinfections was also an undercount. Further, our study population was likely biased toward those with more severe illnesses from initial infection (i.e., those who were hospitalized and thus eligible to access testing during the first wave of infections), which might have resulted in a population with stronger infection-induced immunity [26]. Second, some NYC residents were more likely to be repeatedly tested (e.g., as an occupational requirement particularly if unvaccinated), increasing the probability of ascertaining mild initial infections and/or reinfections. Further, if vaccinated persons were less likely to be tested, then given disproportionate ascertainment of reinfections among unvaccinated persons, VE could be overestimated. However, by assessing VE for symptomatic reinfection and for reinfection with hospitalization, we reduced this potential ascertainment bias from discretionary testing for non-clinical reasons. Third, exposure misclassification is possible because persons vaccinated outside of New York State and by federal programs were likely misclassified as unvaccinated; however, we expect any such misclassification was nondifferential by outcome status. Further, NYC residents who tested positive for SARS-CoV-2 infection in 2020 and then moved out-of-jurisdiction and were reinfected could have been misclassified as control subjects; however, we expect any such misclassification was nondifferential by vaccination status. In addition, some persons with persistent positivity from their first infection in 2020 [27] could have been misclassified as reinfected, although this is unlikely given the minimum 5.5-month interval between the date of first positive test in 2020 and the study period beginning June 15, 2021 for first possible date of reinfection. Also, because reason for hospitalization was not always known, some patients who met COVID-19 hospitalization criteria could have been admitted for another reason unrelated to COVID-19 (e.g., surgery, labor and delivery, or mental health). Finally, while we found that vaccines from all three manufacturers significantly reduced the odds of reinfection, differences in the timing of vaccine availability in the context of potentially waning infection-induced and vaccine-induced immunity and differences in the health status of recipients made direct comparisons between vaccines difficult. In particular, Janssen vaccine did not become available until early March 2021 and was prioritized for more vulnerable groups in NYC, such as homebound seniors and the homeless population, given the operational benefit of requiring only one dose [28].

In summary, our findings counter common reasons for COVID-19 vaccination hesitancy, including the belief that vaccines are unnecessary because of immunity from prior infection or that vaccines might not be protective against new variants. Among adult NYC residents with documented infection in 2020, a small percentage (0.3%) were reinfected during a 2.5-month period when Delta predominated, but unvaccinated versus fully vaccinated individuals had elevated odds of SARS-CoV-2 reinfection and of reinfection with hospitalization. Vaccines from all three manufacturers were similarly and significantly effective. These results reinforce recommendations to vaccinate eligible persons, even if previously infected and especially while the Delta variant predominates.

## Supporting information

Strobe checklist: case-control studies

## Data Availability

Line-level data are not publicly available in accordance with patient confidentiality and privacy laws. Publicly available data are linked below.

https://www1.nyc.gov/site/doh/covid/covid-19-data.page

## Funding

The authors received no specific funding for this work beyond their usual salaries. ALR was supported by ELC CARES (grant No. NU50CK000517-01-09), funded by the US Centers for Disease Control and Prevention (CDC). LF was supported by the ELC COVID Data Analysis Project (BCD) (grant No. 4021-04Z), funded by CDC. SKG was supported by the Public Health Emergency Preparedness Cooperative Agreement (grant No. NU90TP922035-03-03), funded by CDC.

## Acknowledgments

The authors thank all DOHMH staff serving in the Surveillance and Epidemiology Section and Vaccine Operations Center of the Incident Command System, including Jennifer Baumgartner, Katelynn Devinney, Daniel Bertolino, and Dr. Alexandra Ternier for contributions to data management, Dr. Corinne Thompson for constructive manuscript review, and Matthew Montesano for contributions to data communication.

## Supplementary Material

In a matched case-control study, confounding might not be controlled for when estimating the association between exposure and outcome because matching cases with controls by covariates does not necessarily make exposed and unexposed groups balanced [1, 2]. In addition to potential confounding bias, a matched case-control study might be biased due to unequal selection probabilities between cases and controls [2]. To detect and address potential bias in this matched case-control study, we performed a sensitivity analysis where the original matched case-control data were re-analyzed using the case-control weighted targeted maximum likelihood estimation method (CCW-TMLE) [2, 3]. This causal inference method can address selection bias via weighting and confounding bias and model misspecification via doubly robust estimation and machine learning [2-4].

Using the same definitions of case-patients as individuals who tested positive for SARS-CoV-2 reinfection during June 15–August 31, 2021, control subjects, and variables as described in the Methods and with fully vaccinated persons as the referent, we developed models for the unvaccinated (**Model 1**) and for the partially vaccinated (**Model 2**), and re-analyzed the same data from the original conditional logistic regression using CCW-TMLE. We first calculated case- and control-specific weights (**Model 1**: cases = 1, controls = 1/(2,853/349,598); **Model 2**: cases = 1, controls = 1/(1,753/349,598)), where 349,598 was the number of adults who tested positive for SARS-CoV-2 infection in 2020, did not have another positive test >90 days after initial positive test, and did not die before June 15, 2021. We then estimated risk ratios (RR), odds ratios (OR), and their corresponding 95% confidence intervals (CI) using the following procedures for both **Models 1** and **2**. Note that both models used the same analytic steps except for different weights and exposure definitions. The outcome model for reinfection was constructed using five covariates (age group [18–49, 50–64, ≥65 years]; gender; four-level census tract-based poverty level; month of initial positive SARS-CoV-2 test in 2020; and, because living in communities with lower vaccination rates could serve as a common cause of exposure to both vaccination and reinfection [5], the percentage of residents in an individual’s modified ZIP Code tabulation area of residence who were fully vaccinated as of June 15, 2021. This model in turn produced weighted conditional expectations of reinfection under the actual vaccination condition as well as counterfactuals for each person. An additional model for vaccination (propensity score) was constructed using the same set of covariates from the outcome model. Weighted likelihood of receiving an exposure (propensity score; unvaccinated or partially vaccinated versus fully vaccinated) was explicitly estimated and then incorporated values of the outcome variable predicted by the outcome model. This process helps address bias due to differences in these five covariates by unvaccinated or partially vaccinated versus fully vaccinated (propensity score). In addition, use of the same set of variables in the outcome and propensity models helps reduce bias due to misspecification [6, 7]. Estimation was made using the machine learning approach via Superlearner [8]. Specifically, the approach uses a data-adaptive algorithm whereby a series of estimators were calculated via various methods such as random forest, elastic net, regression trees, generalized linear model, generalized linear model with pairwise variable interactions, and generalized linear model with stepwise model selection, and the best weighted combination of estimators are selected via cross-validation, which could address bias due to model misspecification and other violations of statistical assumptions.

Inference was made by the efficient influence curve equation. TMLE analyses were conducted using R software LTMLE package [9]. Statistical significance was determined by a 2-sided p-value <0.05.

Supplementary Table 1 shows that being not vaccinated versus fully vaccinated was associated with 2.20 times the risk of reinfection (95% CI = 1.88, 2.58) and 2.21 times the odds of reinfection (95% CI = 1.89, 2.58). Being partially vaccinated versus fully vaccinated was associated with 1.57 times the risk of reinfection (95% CI = 1.21, 2.03) and 1.57 times the odds of reinfection (95% CI = 1.21, 2.04).

These results are very similar to the results from the conditional logistic regression analysis (Table 2) where mOR for reinfection comparing not vaccinated versus fully vaccinated was 2.23 (95% CI = 1.90, 2.61) and mOR comparing partially vaccinated vs. fully vaccinated was 1.58 (95% CI = 1.22, 2.06). Thus, bias in the matched case-control study was likely negligible, as a more rigorous causal inference method produces similar results.

**Supplementary Table 1.** Associations by using case-control weighted targeted maximum likelihood estimation method between SARS-CoV-2 reinfection during June 15–August 31, 2021 and COVID-19 vaccination status among New York City adults with first SARS-CoV-2 infection in 2020

|  | Risk ratio for reinfection<br>(95% confidence interval) | Odds ratio for<br>reinfection<br>(95% confidence<br>interval) |
| --- | --- | --- |
| Not vaccinated versus fully<br>vaccinated | 2.20 (1.88, 2.58) | 2.21 (1.89, 2.58) |
| Partially vaccinated versus fully<br>vaccinated | 1.57 (1.21, 2.03) | 1.57 (1.21, 2.04) |

## Footnotes

1 Estimates were potentially unreliable (relative standard error >30% or 95% confidence interval half-width >10) and should be interpreted with caution.

## Notes

### Competing Interest Statement

The authors have declared no competing interest.

### Author Declarations

This work was deemed public health surveillance that is non-research by the Department of Health and Mental Hygiene Institutional Review Board.

## References

1. Thompson CN, Baumgartner J, Pichardo C, et al. COVID-19 Outbreak - New York City, February 29-June 1, 2020. MMWR Morb Mortal Wkly Rep 2020; 69(46): 1725–9.

2. New York City Department of Health and Mental Hygiene. COVID-19: Data. Available at: https://www1.nyc.gov/site/doh/covid/covid-19-data.page. Accessed October 19, 2021.

3. New York City Department of Health and Mental Hygiene. 2020 Advisory #8: COVID- 19 Update for New York City. Available at: https://www1.nyc.gov/assets/doh/downloads/pdf/han/advisory/2020/covid-19-03202020.pdf. Accessed October 18, 2021.

4. New York State Governor’s Office. Video, Audio, Photos & Rush Transcript: Governor Cuomo Announces First Dose of COVID-19 Vaccine Administered in United States. Available at: https://www.governor.ny.gov/news/video-audio-photos-rush-transcript-governor-cuomo-announces-first-dose-covid-19-vaccine. Accessed September 28, 2021.

5. Scobie HM, Johnson AG, Suthar AB, et al. Monitoring Incidence of COVID-19 Cases, Hospitalizations, and Deaths, by Vaccination Status - 13 U.S. Jurisdictions, April 4-July 17, 2021. MMWR Morb Mortal Wkly Rep 2021; 70(37): 1284–90.

6. Thompson MG, Burgess JL, Naleway AL, et al. Prevention and Attenuation of Covid-19 with the BNT162b2 and mRNA-1273 Vaccines. N Engl J Med 2021; 385(4): 320–9.

7. Tenforde MW, Self WH, Naioti EA, et al. Sustained Effectiveness of Pfizer-BioNTech and Moderna Vaccines Against COVID-19 Associated Hospitalizations Among Adults - United States, March-July 2021. MMWR Morb Mortal Wkly Rep 2021; 70(34): 1156–62.

8. Haas EJ, Angulo FJ, McLaughlin JM, et al. Impact and effectiveness of mRNA BNT162b2 vaccine against SARS-CoV-2 infections and COVID-19 cases, hospitalisations, and deaths following a nationwide vaccination campaign in Israel: an observational study using national surveillance data. Lancet 2021; 397(10287): 1819–29.

9. Bernal JL, Andrews N, Gower C, et al. Effectiveness of Covid-19 Vaccines against the B.1.617.2 (Delta) Variant. New England Journal of Medicine 2021; 385(7): 585–94.

10. Gee J, Marquez P, Su J, et al. First Month of COVID-19 Vaccine Safety Monitoring - United States, December 14, 2020-January 13, 2021. MMWR Morb Mortal Wkly Rep 2021; 70(8): 283–8.

11. Shay DK, Gee J, Su JR, et al. Safety Monitoring of the Janssen (Johnson & Johnson) COVID-19 Vaccine - United States, March-April 2021. MMWR Morb Mortal Wkly Rep 2021; 70(18): 680–4.

12. New York State Governor’s Office. Governor Cuomo Announces COVID-19 Vaccination Mandate for Healthcare Workers. Available at: https://www.governor.ny.gov/news/governor-cuomo-announces-covid-19-vaccination-mandate-healthcare-workers. Accessed September 28, 2021.

13. The City of New York - Office of the Mayor. Executive Order No. 78: Mandatory Vaccination or Test Requirement for City Employees and Covered Employees of City Contractors. Available at: https://www1.nyc.gov/assets/home/downloads/pdf/executive-orders/2021/eo-78.pdf. Accessed September 28, 2021.

14. The City of New York - Office of the Mayor. Emergency Executive Order No. 225: Key to NYC: Requiring COVID-19 Vaccination for Indoor Entertainment, Recreation, Dining and Fitness Settings. Available at: https://www1.nyc.gov/assets/doh/downloads/pdf/covid/covid-19-key-to-nyc-executive-order.pdf. Accessed September 28, 2021.

15. Ruiz JB, Bell RA. Predictors of intention to vaccinate against COVID-19: Results of a nationwide survey. Vaccine 2021; 39(7): 1080–6.

16. World Health Organization. Tracking SARS-CoV-2 variants. Available at: https://www.who.int/en/activities/tracking-SARS-CoV-2-variants/. Accessed September 28, 2021.

17. Harder T, Kulper-Schiek W, Reda S, et al. Effectiveness of COVID-19 vaccines against SARS-CoV-2 infection with the Delta (B.1.617.2) variant: second interim results of a living systematic review and meta-analysis, 1 January to 25 August 2021. Euro Surveill 2021; 26(41).

18. Planas D, Veyer D, Baidaliuk A, et al. Reduced sensitivity of SARS-CoV-2 variant Delta to antibody neutralization. Nature 2021; 596(7871): 276–80.

19. Cavanaugh AM, Spicer KB, Thoroughman D, Glick C, Winter K. Reduced Risk of Reinfection with SARS-CoV-2 After COVID-19 Vaccination - Kentucky, May-June 2021. MMWR Morb Mortal Wkly Rep 2021; 70(32): 1081–3.

20. United States Centers for Disease Control and Prevention. COVID Data Tracker: United States - Variant Proportions. Available at: https://covid.cdc.gov/covid-data-tracker/#variant-proportions. Accessed September 28, 2021.

21. United States Centers for Disease Control and Prevention. National Notifiable Diseases Surveillance System (NNDSS): Coronavirus Disease 2019 (COVID-19). Available at: https://ndc.services.cdc.gov/conditions/coronavirus-disease-2019-covid-19/. Accessed September 28, 2021.

22. United States Census Bureau. 2014-2018 American Community Survey 5-year Estimates. In: Bureau USC. Washington, D.C., 2018.

23. New York City Department of Health and Mental Hygiene. Citywide Immunization Registry (CIR). Available at: https://www1.nyc.gov/site/doh/providers/reporting-and-services/citywide-immunization-registry-cir.page. Accessed September 28, 2021.

24. Self WH, Tenforde MW, Rhoads JP, et al. Comparative Effectiveness of Moderna, Pfizer-BioNTech, and Janssen (Johnson & Johnson) Vaccines in Preventing COVID-19 Hospitalizations Among Adults Without Immunocompromising Conditions - United States, March-August 2021. MMWR Morb Mortal Wkly Rep 2021; 70(38): 1337–43.

25. Centers for Disease Control and Prevention. Preparing for Your COVID-19 Vaccination. Available at: https://www.cdc.gov/coronavirus/2019-ncov/vaccines/prepare-for-vaccination.html. Accessed November 29, 2021.

26. Lynch KL, Whitman JD, Lacanienta NP, et al. Magnitude and Kinetics of Anti-Severe Acute Respiratory Syndrome Coronavirus 2 Antibody Responses and Their Relationship to Disease Severity. Clin Infect Dis 2021; 72(2): 301–8.

27. Cento V, Colagrossi L, Nava A, et al. Persistent positivity and fluctuations of SARS-CoV-2 RNA in clinically-recovered COVID-19 patients. J Infect 2020; 81(3): e90–e2.

28. The City of New York - Office of the Mayor. Vaccine For All: City Begins Vaccination for Homebound New Yorkers. Available at: https://www1.nyc.gov/office-of-the-mayor/news/155-21/vaccine-all-city-begins-vaccination-homebound-new-yorkers. Accessed September 28, 2021.

## Supplementary References

1. Mansournia MA, Hernán MA, Greenland S. Matched designs and causal diagrams. International journal of epidemiology. 2013;42(3):860–9.

2. Rose S, van der Laan M. A double robust approach to causal effects in case-control studies. American journal of epidemiology. 2014;179(6):663–9.

3. Almasi-Hashiani A, Nedjat S, Ghiasvand R, Safiri S, Nazemipour M, Mansournia N, Mansournia MA. The causal effect and impact of reproductive factors on breast cancer using super learner and targeted maximum likelihood estimation: a case-control study in Fars Province, Iran. BMC Public Health. 2021;21(1):1–8.

4. Van der Laan MJ. Targeted maximum likelihood based causal inference: Part I. International Journal of Biostatistics. 2010;6(2).

5. Siegel DA, Reses HE, Cool AJ, et al. Trends in COVID-19 Cases, Emergency Department Visits, and Hospital Admissions Among Children and Adolescents Aged 0–17 Years — United States, August 2020–August 2021. MMWR Morbidity and Mortality Weekly Report 2021;70(36):1249–54.

6. Bang H, Robins JM. Doubly robust estimation in missing data and causal inference models. Biometrics. 2005;61(4):962–73.

7. Petersen M, Schwab J, Gruber S et al. Targeted maximum likelihood estimation for dynamic and static longitudinal marginal structural working models. Journal of Causal Inference. 2014;2(2):147–85.

8. van der Laan MJ, Polley EC, Hubbard AE. Super Learner. Statistical Applications in Genetics and Molecular Biology. 2007;6(1), 1–21.

9. Lendle S, Schwab J, Petersen ML, van der Laan MJ. ltmle: an R package implementing targeted minimum loss-based estimation for longitudinal data. Journal of Statistical Software. 2017;81(1):1–21.

